# Forecasting COVID-19 pandemic Severity in Asia

**DOI:** 10.1101/2020.05.15.20102640

**Authors:** Elinor Aviv-Sharon, Asaph Aharoni

**Affiliations:** Department of Plant and Environmental Sciences, Weizmann Institute of Science, Rehovot, 7610001, Israel

**Keywords:** COVID-19, SARS-CoV-2, emerging infectious diseases, forecasting, prediction, generalized logistic model, Richards model, epidemic modeling

## Abstract

Four months into the ongoing novel coronavirus disease 2019 (COVID-19) pandemic, this work provides a simple and direct projection of the outbreak spreading potential and the pandemic cessation dates in China, Iran, the Philippines and Taiwan, using the generalized logistic model (GLM). The short-term predicted number of cumulative COVID-19 cases matched the confirmed reports of those who were infected across the four countries, suggesting GLM as a valuable tool for characterizing the transmission dynamics process and the trajectory of COVID-19 pandemic along with the impact of interventions.

## Introduction

Coronavirus disease 2019 (COVID-19), caused by the novel severe acute respiratory syndrome coronavirus 2 (SARS-CoV-2) was identified in China, in November 2019, declared to be a public health emergency of international concern in two months, and recognized as a pandemic on 11^th^ March 2020. As of 10^th^ April 2020, approximately 1.7 million cases of COVID-19 have been reported in 210 countries and territories, resulting in approximately 95,800 deaths. Emerging infectious diseases (EID), which appear for the first time, or that may have existed previously but is rapidly are possibly the deadliest and continue to challenge human health.

As the world races to find a vaccine or a treatment to combat the pandemic, many concerns arise about the outbreak severity, particularly the potential number of infected people. Hence, it is of a great importance to estimate the outbreak evolution using epidemiology models. Here, the epidemiological dataset of confirmed cases with COVID-19 in China, Iran, the Philippines and Taiwan, as of 10^th^ April 2020, was analyzed, using the generalized logistic model (GLM), also known as Richards’ model. This empirical function has made many remarkable coincidences with real SARS, Zika and Ebola epidemic data for real-time prediction of outbreak development *(1–4)*. Early assessment of the severity of infection and transmissibility can help quantify COVID-19 pandemic potential and anticipate the likely number of infected people by the end of the epidemics.

## Data and methods

Specific countries were selected as denoting different COVID-19 incidence scales. China and Iran, the two major centers of COVID-19 outbreak in eastern and southern Asia, respectively with tens of thousands of cases; the Philippines, a representative of an archipelagic country with thousands of cases, and Taiwan, with only hundreds of cases. For each country, the officially reported data on COVID-19 daily cases from the onset of the outbreak to April 10^th^, 2020 were collected from governmental or health authorities’ websites (Appendix Table, Figure). To allow outbreak projection, the data from the early phase of the outbreak (35–40 days) were fitted with the GLM as described previously *(1,5)*. Briefly, this approach enables the evaluation of the cumulative number of COVID-19 cases, represented by *Y(t)*. The dynamics of *Y*, in period *t*, can be expressed as:

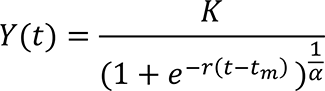

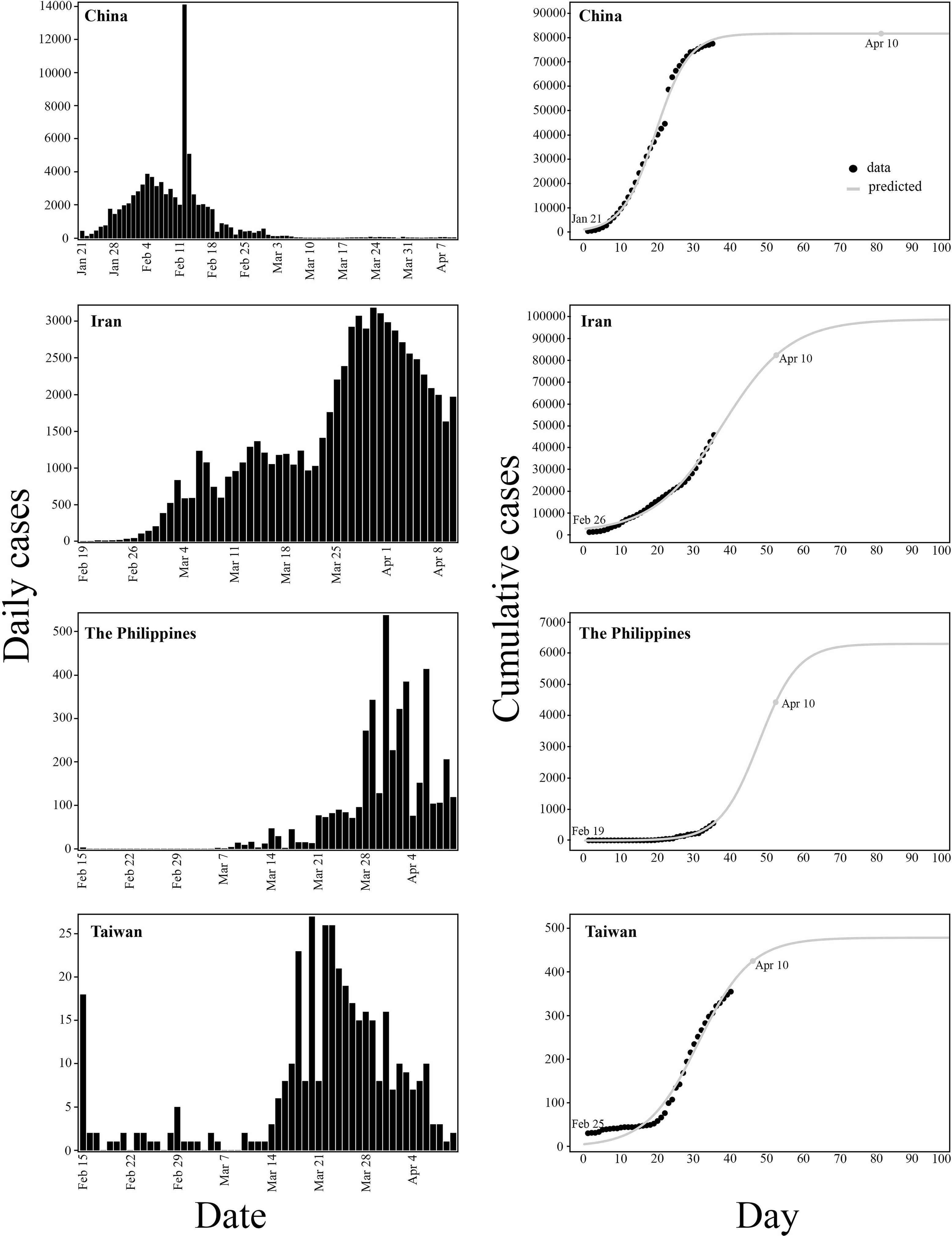
The generalized logistic growth model-predicted size of the COVID-19 pandemic in China, Iran, the Philippines and Taiwan. On the left: the daily number of new confirmed COVID-19 cases. On the right: the observed (black circles) and the model-fitted and predicted cumulative cases (grey solid line) over time. The grey circle denotes the predicted number of cumulative cases as of 10 April 2020.

Where *K* is the upper asymptote, or the maximum cumulative case incidence, *r* is the intrinsic growth rate and *tm* is the turning point, the time where maximum number of cases per day occur. The model predicts that cumulative COVID-19 case incidence follows an S-shaped curve and gradually reaches *K*. Not a single new COVID-19 case emerging within 3 consecutive months defines the end of the epidemic *(1)*.

The basic reproduction number of COVID-19, R_0_, the expected number of secondary cases produced by a single infection was estimated as *R*_0_ = *e*^*rT*^, where T is the mean serial interval; the time that elapses between onset of symptoms in the primary case and onset of symptoms of the secondary case.

The model should conform to several assumptions: first, as the number of tests conducted affects the reported number of daily cases, similar number of individuals are tested daily. Secondly, cases are not imported from outside the country. Thirdly, the model does not consider human behavior and is conditional on the assumption that public gatherings are highly limited, allowing the epidemic to follow its natural course. Likewise, implementation of intervention measures, such as enhanced hygiene, isolation, contact tracing, restrictions on social contacts and migration by air or train, are maintained continuously.

## Results and Discussion

Corresponding to the basic premise of GLM, the cumulative COVID-19 cases curve of each country consists of a single peak of high incidence, resulting in a sigmoid curve with a single turning point (Figure). For all localities, high correlations between observed and predicted incidence were found (R^2^>0.99, p value<2.2e^−16^). To evaluate the forecasting performance of the model, the total number of cases on April 10^th^ were estimated based on the observed incidence during the initial stage of the pandemic. A subset data of 35 days was used for China (January 21^st^ to February 24^th^ 2020), Iran (February 26^th^ to March 31^st^ 2020) and the Philippines (February 19^th^ to March 24^th^ 2020). In Taiwan, due to a smaller scale of cases, dataset of 40 days (February 25^th^ to April 4^th^ 2020) was required for optimal fit to the model. Forecasting the total number of cases on April 10^th^ 2020 (Figure), was done 46, 10, 17 and 6 consecutive days ahead for China, Iran, the Philippines and Taiwan, respectively. In all cases, the predicted cumulative number of cases was similar to the one officially reported (Table). The predicted number was 81797 for China, 68225 for Iran, 4430 for the Philippines and 425 for Taiwan, while the observed total number of cases was 81953, 68192, 4195 and 382, respectively. The maximum predicted cumulative incidence, *K*, was estimated to be 81797 for China, 97576 for Iran, 6300 for the Philippines, and 479 for Taiwan [see Table for 95% confidence interval (CI)]. The earliest time for the current COVID-19 pandemic to cease, was evaluated to occur after 73 days (April 2^nd^) in China, 147 days (July 21^st^) in Iran, 122 days (June 19^th^) in the Philippines, and 80 days (May 14^th^) in Taiwan. A mean serial interval of 5.8 days *(6)* was used to calculate R0, the basic reproduction number of COVID-19 infections. R_0_ estimates were 3.59, 1.86, 2.99 and 2.26 in China, Iran, the Philippines and Taiwan, respectively, similar to those published previously *(7)* and to that of SARS *(8)*. These early stages R0 are likely to decrease with control measurements policies continuation. Indeed, predicting COVID-19 dynamics based on its initial growth phase, revealed that the turning point of each country has occurred closer to the lower limit of the 95% CI and even earlier than expected, due to the control measurements effectiveness. The turning point was estimated as day 19.4±0.27 in China, 37.1±3.6 in Iran, 47.4±11.6 in the Philippines and 31.2±1.5 in Taiwan, whereas the observed highest number of daily cases in China (14108), Iran (3186), the Philippines (538) and Taiwan (27) has occurred on day 23, 34, 42, and 23, respectively (Table).

**Table 1:** The generalized logistic growth model estimates of COVID-19 pandemic

**Table. The generalized logistic growth model estimates of COVID-19 pandemic**
| Country | $R^{2*}$ | $r$ | $R_0$ | $t_m$<br>projected | $t_m$ observed | Maximum<br>No. of cases<br>(95% CI<sup>†</sup>) | Pandemic<br>cessation<br>date | 10 April<br>projected<br>No. of<br>cases | 10 April<br>observed<br>No. of<br>cases |
| --- | --- | --- | --- | --- | --- | --- | --- | --- | --- |
| China | 0.997 | 0.22 | 3.59 | 19.4±0.27 | Day 23<br>(February<br>12<sup>th</sup>) | 81797±1445 | Day 73<br>(April 2<sup>nd</sup>) | 81797 | 81953 |
| Iran | 0.995 | 0.1 | 1.86 | 37.1±3.6 | Day 34<br>(March 30<sup>th</sup>) | 97576±22098 | Day 147<br>(July 21<sup>st</sup>) | 68225 | 68192 |
| The<br>Philippines | 0.995 | 0.18 | 2.99 | 47.4±11.6 | Day 42<br>(March 31<sup>st</sup>) | 6300±11630 | day 122<br>(June 19<sup>th</sup>) | 4430 | 4195 |
| Taiwan | 0.99 | 0.14 | 2.26 | 31.2±1.5 | Day 23<br>(March 20<sup>th</sup>) | 479±44 | day 80<br>(May 14<sup>th</sup>) | 425 | 382 |

Forecasting COVID-19 pandemic is challenging in the context of an outbreak caused by novel pathogen for which its natural history and modes of transmission are unknown. Since the GLM is trained on the existing data and is designed to fit the development of epidemic curves, rather than EID estimation only, it could provide a good fit to the limited available COVID-19 epidemiological data to characterize the transmission dynamics process and the trajectory of COVID-19 pandemic along with the impact of interventions *(9)*. This prediction is conditional to intervention measures continuation. Changes of the current policies or human behavior may affect the actual contact rate and the subsequent development of the epidemic. Additionally, testing kits deficiency can lead to poor diagnosis and incomplete data, which may implicate the model robustness. Even so, forecasting epidemic size and peak time may help clarify what the future holds, and could be useful to make long-term strategic decisions regarding the distribution of testing and treatment facilities that may be required, and may be helpful to assess the extent of protective and medical al equipment needed for the near future.

## Data Availability

See Appendix Table

## Acknowledgments

We thank the Adelis Foundation, Leona M. and Harry B. Helmsley Charitable Trust, Jeanne and Joseph Nissim Foundation for Life Sciences, Tom and Sondra Rykoff Family Foundation Research and the Raymond Burton Plant Genome Research Fund for supporting the AA lab activity. A.A. is the incumbent of the Peter J. Cohn Professorial Chair. The authors report no funding related to this research and no competing interests.

**Appendix Table.**
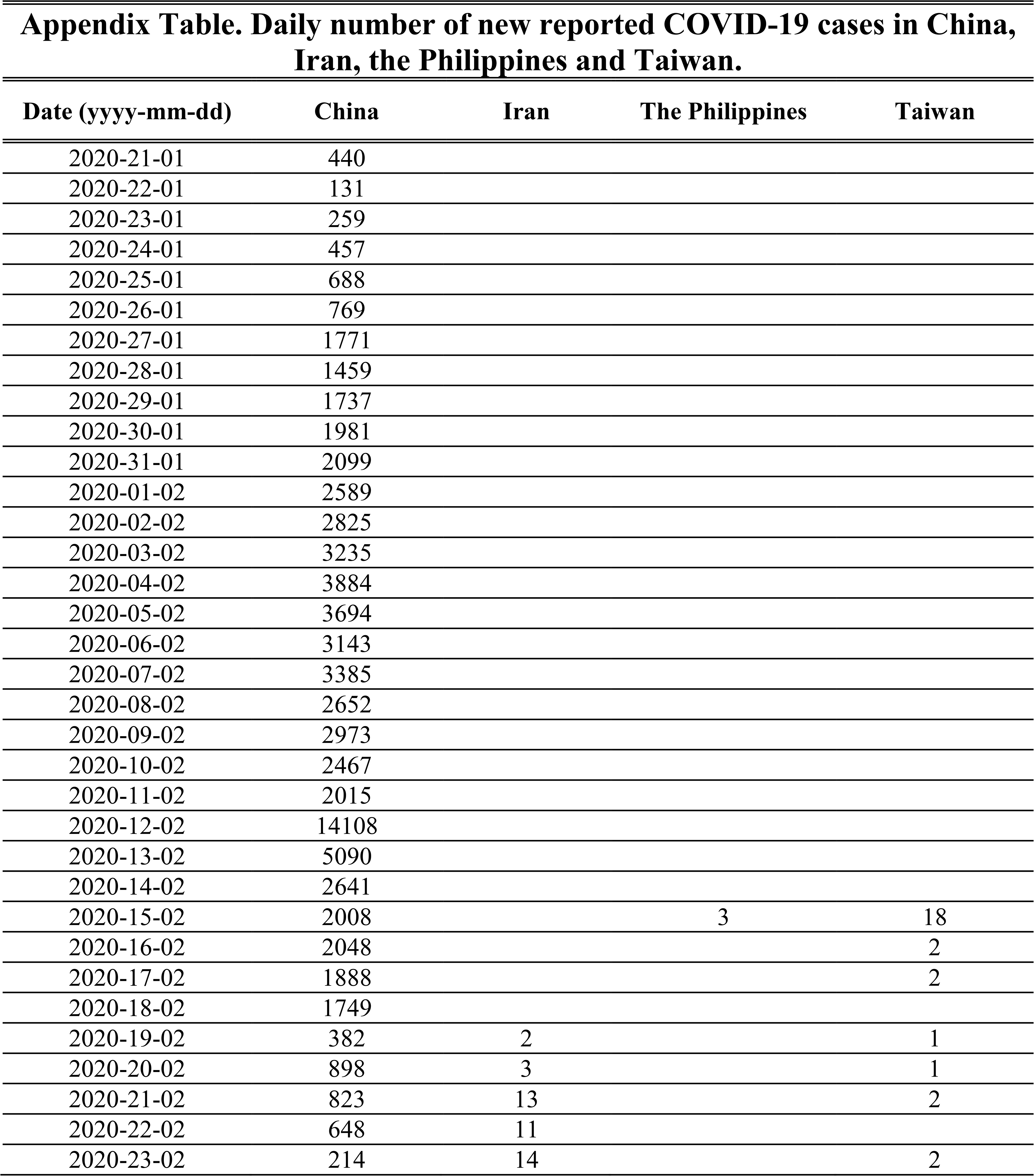
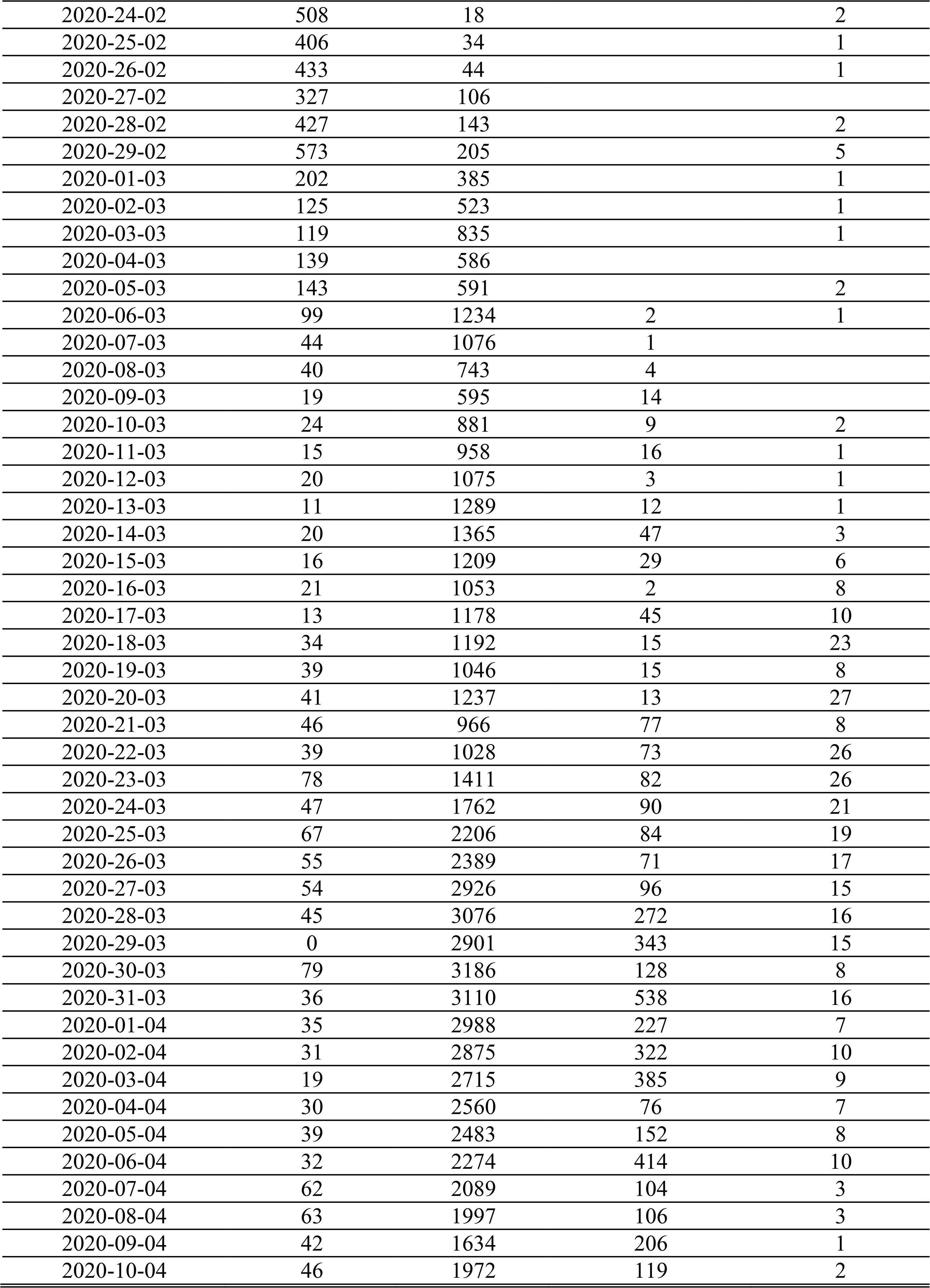
Daily number of new reported COVID-19 cases in China, Iran, the Philippines and Taiwan.

| Date (yyyy-mm-dd) | China | Iran | The Philippines | Taiwan |
| --- | --- | --- | --- | --- |
| 2020-21-01 | 440 |  |  |  |
| 2020-22-01 | 131 |  |  |  |
| 2020-23-01 | 259 |  |  |  |
| 2020-24-01 | 457 |  |  |  |
| 2020-25-01 | 688 |  |  |  |
| 2020-26-01 | 769 |  |  |  |
| 2020-27-01 | 1771 |  |  |  |
| 2020-28-01 | 1459 |  |  |  |
| 2020-29-01 | 1737 |  |  |  |
| 2020-30-01 | 1981 |  |  |  |
| 2020-31-01 | 2099 |  |  |  |
| 2020-01-02 | 2589 |  |  |  |
| 2020-02-02 | 2825 |  |  |  |
| 2020-03-02 | 3235 |  |  |  |
| 2020-04-02 | 3884 |  |  |  |
| 2020-05-02 | 3694 |  |  |  |
| 2020-06-02 | 3143 |  |  |  |
| 2020-07-02 | 3385 |  |  |  |
| 2020-08-02 | 2652 |  |  |  |
| 2020-09-02 | 2973 |  |  |  |
| 2020-10-02 | 2467 |  |  |  |
| 2020-11-02 | 2015 |  |  |  |
| 2020-12-02 | 14108 |  |  |  |
| 2020-13-02 | 5090 |  |  |  |
| 2020-14-02 | 2641 |  |  |  |
| 2020-15-02 | 2008 |  | 3 | 18 |
| 2020-16-02 | 2048 |  |  | 2 |
| 2020-17-02 | 1888 |  |  | 2 |
| 2020-18-02 | 1749 |  |  |  |
| 2020-19-02 | 382 | 2 |  | 1 |
| 2020-20-02 | 898 | 3 |  | 1 |
| 2020-21-02 | 823 | 13 |  | 2 |
| 2020-22-02 | 648 | 11 |  |  |
| 2020-23-02 | 214 | 14 |  | 2 |
| 2020-24-02 | 508 | 18 |  | 2 |
| 2020-25-02 | 406 | 34 |  | 1 |
| 2020-26-02 | 433 | 44 |  | 1 |
| 2020-27-02 | 327 | 106 |  |  |
| 2020-28-02 | 427 | 143 |  | 2 |
| 2020-29-02 | 573 | 205 |  | 5 |
| 2020-01-03 | 202 | 385 |  | 1 |
| 2020-02-03 | 125 | 523 |  | 1 |
| 2020-03-03 | 119 | 835 |  | 1 |
| 2020-04-03 | 139 | 586 |  |  |
| 2020-05-03 | 143 | 591 |  | 2 |
| 2020-06-03 | 99 | 1234 | 2 | 1 |
| 2020-07-03 | 44 | 1076 | 1 |  |
| 2020-08-03 | 40 | 743 | 4 |  |
| 2020-09-03 | 19 | 595 | 14 |  |
| 2020-10-03 | 24 | 881 | 9 | 2 |
| 2020-11-03 | 15 | 958 | 16 | 1 |
| 2020-12-03 | 20 | 1075 | 3 | 1 |
| 2020-13-03 | 11 | 1289 | 12 | 1 |
| 2020-14-03 | 20 | 1365 | 47 | 3 |
| 2020-15-03 | 16 | 1209 | 29 | 6 |
| 2020-16-03 | 21 | 1053 | 2 | 8 |
| 2020-17-03 | 13 | 1178 | 45 | 10 |
| 2020-18-03 | 34 | 1192 | 15 | 23 |
| 2020-19-03 | 39 | 1046 | 15 | 8 |
| 2020-20-03 | 41 | 1237 | 13 | 27 |
| 2020-21-03 | 46 | 966 | 77 | 8 |
| 2020-22-03 | 39 | 1028 | 73 | 26 |
| 2020-23-03 | 78 | 1411 | 82 | 26 |
| 2020-24-03 | 47 | 1762 | 90 | 21 |
| 2020-25-03 | 67 | 2206 | 84 | 19 |
| 2020-26-03 | 55 | 2389 | 71 | 17 |
| 2020-27-03 | 54 | 2926 | 96 | 15 |
| 2020-28-03 | 45 | 3076 | 272 | 16 |
| 2020-29-03 | 0 | 2901 | 343 | 15 |
| 2020-30-03 | 79 | 3186 | 128 | 8 |
| 2020-31-03 | 36 | 3110 | 538 | 16 |
| 2020-01-04 | 35 | 2988 | 227 | 7 |
| 2020-02-04 | 31 | 2875 | 322 | 10 |
| 2020-03-04 | 19 | 2715 | 385 | 9 |
| 2020-04-04 | 30 | 2560 | 76 | 7 |
| 2020-05-04 | 39 | 2483 | 152 | 8 |
| 2020-06-04 | 32 | 2274 | 414 | 10 |
| 2020-07-04 | 62 | 2089 | 104 | 3 |
| 2020-08-04 | 63 | 1997 | 106 | 3 |
| 2020-09-04 | 42 | 1634 | 206 | 1 |
| 2020-10-04 | 46 | 1972 | 119 | 2 |

## Notes

### Competing Interest Statement

The authors have declared no competing interest.

